# Evaluating the contributions of strategies to prevent SARS-CoV-2 transmission in the healthcare setting: a modelling study

**DOI:** 10.1101/2020.04.20.20073080

**Authors:** Joel C. Miller, Xueting Qiu, Derek R. MacFadden, William P. Hanage

## Abstract

**Background:** Since its onset, the COVID-19 pandemic has caused significant morbidity and mortality worldwide, with particularly severe outcomes in healthcare institutions and congregate settings. To mitigate spread, healthcare systems have been cohorting patients to limit contacts between uninfected patients and potentially infected patients or healthcare workers (HCWs). A major challenge in managing the pandemic is the presence of currently asymptomatic individuals capable of transmitting the virus, who could introduce COVID-19 into uninfected cohorts. The optimal combination of personal protective equipment (PPE) and testing approaches to prevent these events is unclear, especially in light of ongoing limitations in access to both.

**Methods:** Using stochastic simulations with an SEIR model we quantified and compared the impacts of PPE use, patient and HCWs testing, and cohorting.

**Findings:** In the base case without testing or PPE, the healthcare system was rapidly overwhelmed, and became a net contributor to the force of infection. We found that effective use of PPE by both HCWs and patients could prevent this scenario, while random testing of apparently asymptomatic individuals on a weekly basis was less effective. We also found that even imperfect use of PPE could provide substantial protection by decreasing the force of infection, and that creation of smaller patient/HCW subcohorts can provide additional resilience to outbreak development.

**Interpretation:** These findings reinforce the importance of ensuring adequate PPE supplies even in the absence of testing, and provide support for strict subcohorting regimens to reduce outbreak potential in healthcare institutions.

**Funding:** National Institute of General Medical Sciences, National Institutes of Health.

**Research in context:** *Evidence before:* Preserving healthcare from outbreaks of respiratory viruses is a longstanding concern, brought into sharp relief by the covid-19 pandemic. Early case series and numerous anecdotal reports suggest that health care workers (HCWs) and patients receiving treatment for conditions other than SARS-CoV-2 infection are at elevated risk of becoming infected, and the consequences of infections in long term care facilities are well known. In addition, the early stages of the pandemic have been marked by shortages of personal protective equipment (PPE) and diagnostic testing, but the most effective strategies for their use given the specific characteristics of SARS-CoV-2 transmission are unclear.

*Value added:* Our research plainly shows the importance of presymptomatic transmission. Given reasonable estimates of this, random testing of currently asymptomatic staff and patients once a week is not able to prevent large outbreaks. We show that PPE is, as expected, the most effective intervention and moreover even suboptimal PPE use is highly beneficial. To further limit transmission, we show the benefit of sub-cohorting into smaller groups of HCWs and patients. When the force of infection in the community is low, this can entirely prevent the establishment of infection in a large fraction of healthcare.

*Implications:* PPE should be used throughout healthcare, on the assumption that any patient or HCWs is potentially infected. Further work should determine the most effective means of PPE for the non-COVID cohort. If PPE resources are limited, whether in general or due to a second surge, we recommend subcohorting to limit the impact of introductions from the community.

## Introduction

The COVID-19 pandemic is one the most serious threats to public health in over a century. The number of confirmed infected individuals continues to grow at exponential rates in many countries globally. While comparatively extreme interventions in China, where the pandemic started in 2019, seem to have successfully brought transmission under control,^1^ other locations have yet to achieve this, and may not be able to given local conditions. Even in those regions that have successfully eliminated the pandemic, for now, there remains the risk of re-introduction.

Early reports from Wuhan indicated a relatively large proportion of cases among health care workers (HCWs), who have also been disproportionately represented in confirmed cases from both Italy and the US.^2,3^ Not only is this a major concern for the health of frontline responders, there is also a risk of transmission to patients. For this reason, the design and implementation of cohorting strategies to restrict contact between COVID-19 patients and the rest of the healthcare system are of great importance. Already in some locations, hospitals or other facilities are being exclusively dedicated for COVID-19 patients, separate from others reserved for for the non-COVID-19 cohort, and alternate care facilities are in the process of being established.^4^ China built entirely new hospitals to handle the surge in Wuhan.^1,5^

The use of personal protective equipment (PPE), together with regular testing to identify infected individuals, are important ways of preventing transmission. This is particularly important in healthcare systems throughout the continuum of care, including hospitals, primary care, and long-term care (LTC) facilities such as nursing homes, which have been a focus of the early pandemic. Once SARS-CoV-2 is introduced to these settings, rapid transmission has been observed.^6^ The existence of transmission from asymptomatic or presymptomatic individuals presents a sobering challenge to infection control.^7^ The effective use of PPE – including face masks, face shields, goggles, gloves and gowns – is the major proposed measure to prevent the spread of infection to and from patients and HCWs, but the shortage of many sorts of PPE has already proven a critical problem in many healthcare institutions^8^ and there is no reason to think this will change in the event of future waves of the pandemic. It is often suggested that cohorting low-risk patients is a way to preserve scarce PPE, but for this to be effective and safe requires an extremely high rate of accurate rapid virological testing to detect new infections and prevent further transmission. Such testing regimens are not currently implemented in many settings in the US^8^ or worldwide. With the reality of supply shortage, the optimal combination of viral testing, PPE, and cohort size has not been quantified.

Determining optimal interventions during epidemics can be challenging, as there is often limited time and resources for launching large prospective studies. Moreover, informed decisions about infection control practices need to be made early on. Dynamic models offer a scientific framework with which to predict epidemic outcomes using established parameters and known disease transmission characteristics. Such models have been employed to evaluate potential COVID-19 management approaches,^9^ that have informed physical distancing policy throughout the world. We can use these models to help us better plan PPE use, diagnostic testing approaches, and cohorting.

In this paper we seek to evaluate the impacts of different PPE, diagnostic testing, and patient cohorting strategies in the context of two groups of patients, COVID-19 infected, and COVID-19 uninfected, using a modified SEIR stochastic model. We hypothesized that PPE would be the most effective component of COVID-19 transmission prevention in healthcare, and that subcohorting may add resilience to outbreak development where other interventions are not available. In this paper we explore a cohorting strategy in which the population is divided into the general public and a cohort of HCWs and patients which are kept separate from individuals diagnosed with COVID-19. In practice it may make sense for a community to divide their uninfected patients and HCWs into multiple subcohorts which are intended to contain the impact of an introduction. The dynamics of health care workers and the uninfected patients are stochastic. Within the non-COVID-19 cohort, we assume that once individuals become symptomatic they are identified and removed. We use stochastic simulations to track these components. To demonstrate the impacts of control measures and cohort size, we focus on a single cohort of HCWs and the patients.

## Methods

We consider a basic transmission model. The model has two major components: transmission within the broader community, and transmission within a cohort of HCWs and patients who are initially not infected with SARS-CoV-2, that is, the non-COVID-19 cohort.

We investigate results from a stochastic system to model introductions into the nonCOVID-19 cohort in the healthcare system, assuming that the background dynamics in the community at large follow simple deterministic *SEI*_*A*_*I*_1_*I*_2_*R* dynamics. Individuals begin susceptible (*S*). Following a transmission event, they move into the exposed class (*E*). After that they become infectious. A fraction of these remain asymptomatic (*I*_*A*_) and so can only be identified through testing. Another fraction of infections are initially “pre-sympomatic” (*I*_1_) and eventually exhibit symptoms (*I*_2_) which could lead to identification. Finally they recover with immunity (*R*). We note that we are not modeling the effects of mitigation strategies in this work, but rather the risk of transmission in healthcare settings and the most effective ways to minimize it.

The specific details of our simulations are described in the Appendix. The simulations are scripted in Python (scripts can be found here – https://github.com/joelmiller/HospitalCOVID19). What follows is a broad overview of the model structure. We assume that the broader community follows deterministic dynamics, described by a system of ordinary differential equations.

The HCWs and patient cohort is modelled stochastically using a Gillespie-Doob algorithm.^10^ The cohort experiences introductions either through HCWs infected in the broader community or patients (which may come from visitors or from newly admitted patients who are incorrectly identified as uninfected).

### 1. Variables and Parameters

We will use the variables *S, E, I*_*A*_, *I*_1_, *I*_2_, and *R* for two purposes: both to denote the number of individuals in a particular state, and also as a shorthand to refer to the status of an individual. So the number of *S* individuals in the population is *S*, the number of *E* HCWs in the cohort is *E*_*H*_, and the number of asymptomatically infected patients is *I*_*A,P*_.

Table 1 shows the variables we track with the models, and Tables 2 and 3 show the parameters and their default values.

**Table 1.** The variables used in the model.

| Variable | Definition |
| --- | --- |
| $S, E, I_A, I_1, I_2, R$ | number of susceptible, exposed, asymptomatic infectious, presymptomatic infectious, symptomatic infectious, and recovered individuals in the general population. |
| $S_P, E_P, I_{A,P}, I_{1,P}, R_P$ | number of patients in the cohort. We assume that symptomatic cases are removed immediately. |
| $S_H, E_H, I_{A,H}, I_{1,H}, Q_H, R_H$ | number of HCWs of each status. We assume that identified infections are moved into a quarantine class $Q_H$ until recovering. |
| $N_P = S_P + E_P + I_{A,P} + I_{1,P} + R_P$ and<br>$N_H = S_H + E_H + I_{A,H} + I_{1,H} + R_H$ | Number of patients and health care workers active in the cohort (no symptomatic or quarantined individuals). |

The $I_{2,P}$ , $I_{2,H}$ classes are neglected in the model because we assume individuals are removed as soon as they become symptomatic.

**Table 2.** Default parameter values of disease spread in general population.

| Parameter | Default Value | Definition |
| --- | --- | --- |
| $\lambda_1$ | 1/4 | Average transmission rate from $I_1$ individuals |
| $\lambda_2$ | 2/7 | Average transmission rate from $I_2$ individuals |
| $\lambda_A$ | $\lambda_1$ | Average transmission rate from $I_A$ individuals |
| $\lambda$ | $\lambda_1 I_1 + \lambda_2 I_2 + \lambda_A I_A$ | Overall transmission rate (force of infection) to $S$ individuals in general public. |
| $\gamma_E$ | 1/3 | rate of a transition out of $E$ to either $I_1$ or $I_A$. |
| $q$ | 1/2 | The probability a transition from $E$ is to $I_A$. |
| $\gamma_{I,1}$ | 1/2 | rate of an $I_1 \rightarrow I_2$ transition. |
| $\gamma_{I,2}$ | 1/7 | rate of an $I_2 \rightarrow R$ transition. |
| $\gamma_A$ | 1/9 | The rate of an $I_A \rightarrow R$ transition. |

**Table 3.** The default values for health-care related parameters.

| Parameter | Default Value | Definition |
| --- | --- | --- |
| $\gamma_Q$ | 1/14 | The rate at which quarantined individuals are released. |
| $\omega$ | 0 | Weekly testing rate of HCWs and Patients. |
| $\rho$ | 0 | Probability a non-symptomatic individual would get admitted. |
| $C_H$ | 0.1 | The relative transmission from the general public to HCWs. |
| $C_P$ | 0.1 | The relative transmission from the general public to patients (captures risk from visitors). |
| $C_{PP}$ | 0.5 | Scaling factor for patient to patient transmissions relative to number expected an infected individual would cause in general population. |
| $C_{PH}$ | 2 | Scaling factor for patient to HCW transmission, relative to general population |
| $C_{HP}$ | 2 | Scaling factor for HCW to patient transmission. |
| $C_{HH}$ | 1 | Scaling factor for HCW–HCW transmission within the cohort. |
| $\hat{N}$ | 1000 | the typical size of a cohort in absence of transmission. The natural discharge rate is $b/\hat{N}$. In absence of disease $N_P$ would oscillate around $\hat{N}$. |
| $\hat{N}/4$ | 250 | The total number of HCWs allocated to the cohort (changes when HCWs go into or return from quarantine). |
| $b$ | $\hat{N}/14$ | natural rate at which new patients arrive at a cohort. |

The basic reproduction number in the general population is,

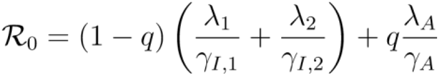

For our values, we find R_0_ = 2·5 − 0·25*q*. If all individuals become symptomatic (*q* = 0) then R_0_ = 2·5, while if all become asymptomatic (*q* = 1) then R_0_ = 2·25. The R_0_ value is in the range of estimates from previous studies.^11–13^ We assume that the average transmission rate in asymptomatic infection is the same as that in pre-symptomatic infection, that is, *λ*_*A*_ = *λ*_1_.

Within the healthcare setting, we expect that HCWs are at high risk of infection, which may be reduced but not eliminated by PPE. This is because of the frequent close interactions between HCWs and their patients. Additionally this expectation is supported by the observed high rates of infection among HCWs in many different populations.^1,3,14,15^ This is reflected in the large value of *C*_*PH*_, representing that an infected patient transmits to HCWs at a rate that is *C*_*PH*_ times that of a general member of the public to other members of the public.

We anticipate that in the absence of intervention to prevent it transmission from HCW to patient will also be relatively high. HCWs transmit to patients at a rate that is *C*_*HP*_ times that of a general member of the public to other members of the public. It should be noted that patients typically outnumber HCWs. So the transmissions from patients to HCWs are concentrated in a smaller population. This means that all else being equal, the force of infection experienced by HCWs is higher than that of patients. So even if *C*_*PH*_ = *C*_*HP*_, this represents a higher transmission probability per interaction from patients to HCWs than *vice versa*.

### 2. Parameter values for different scenarios

To quantify the impacts of different interventions, we defined parameter values for different scenarios in addition to the base model with the default parameter values above, that is, no interventions.

In detail,

- in the scenarios with viral testing, we set the testing rate of patients and HCWs to be weekly i.e. *ω* = 1*/*7.
- To explore the impacts of PPE, several scenarios have been defined:
  − At best, PPE reduces the scaling factor to 1/8 of the default, that is, *C*_*PP*_ = 0·0625, *C*_*PH*_ = 0·25, *C*_*HP*_ = 0.25, and *C*_*HH*_ = 0·125;
  − When only HCWs use PPE, we set *C*_*PP*_ = 0·5 (the default for between-patient transmission), *C*_*PH*_ = 0·25, *C*_*HP*_ = 0·25, and *C*_*HH*_ = 0·125;
  − When both HCWs and patients use PPE, we set *C*_*PP*_ = 0·0625, *C*_*PH*_ = 0·25, *C*_*HP*_ = 0·25, and *C*_*HH*_ = 0·125;
  − The 75% effective PPE is defined as reducing the scaling factor to 1/4 of the default transmissions, that is, *C*_*PP*_ = 0·125, *C*_*PH*_ = 0·5, *C*_*HP*_ = 0·5, and *C*_*HH*_ =0·25;
  − The 50% effective PPE is defined as reducing the scaling factor to half of the default transmissions, that is, *C*_*PP*_ = 0·25, *C*_*PH*_ = 1, *C*_*HP*_ = 1, and *C*_*HH*_ = 0·5.
- To account for the uncertainty of the proportion and the duration of asymptomatic infections, we set
  − lower proportion of asymptomatic infections: q = 0·3; **–** higher proportion of asymptomatic infections: q = 0·7;
  − shorter duration of asymptomatic infections: *γ*_*A*_ = 1*/*5.
- To explore the impacts of cohort size, we run simulations with patient cohort size as 50, 100, 200, 400, 800 and 1600, respectively, with the probability a non-symptomatic individual would get admitted as *ρ* = 0·05.

## Results

### 1. Base model without testing and PPE

We find that in the absence of any attempts to prevent introduction of SARS-CoV-2 to the non-COVID-19 cohort, HCWs rapidly become infected (Figure 1(a)), consistent with general observations from the early stages of the current pandemic.^1,16^ While this leads to a high force of infection to patients in the early stage of the epidemic (Figure 1(a)), later, once many of the HCWs have developed immunity or become symptomatic and moved into quarantine, the force of infection to patients drops. At later stages, as the epidemic grows in the general population, the patients are at reduced risk. This is because the patients primarily interact with HCWs who have been immunized by infection, meanwhile they have relatively little interaction with other patients or the general public.

**Figure 1.**
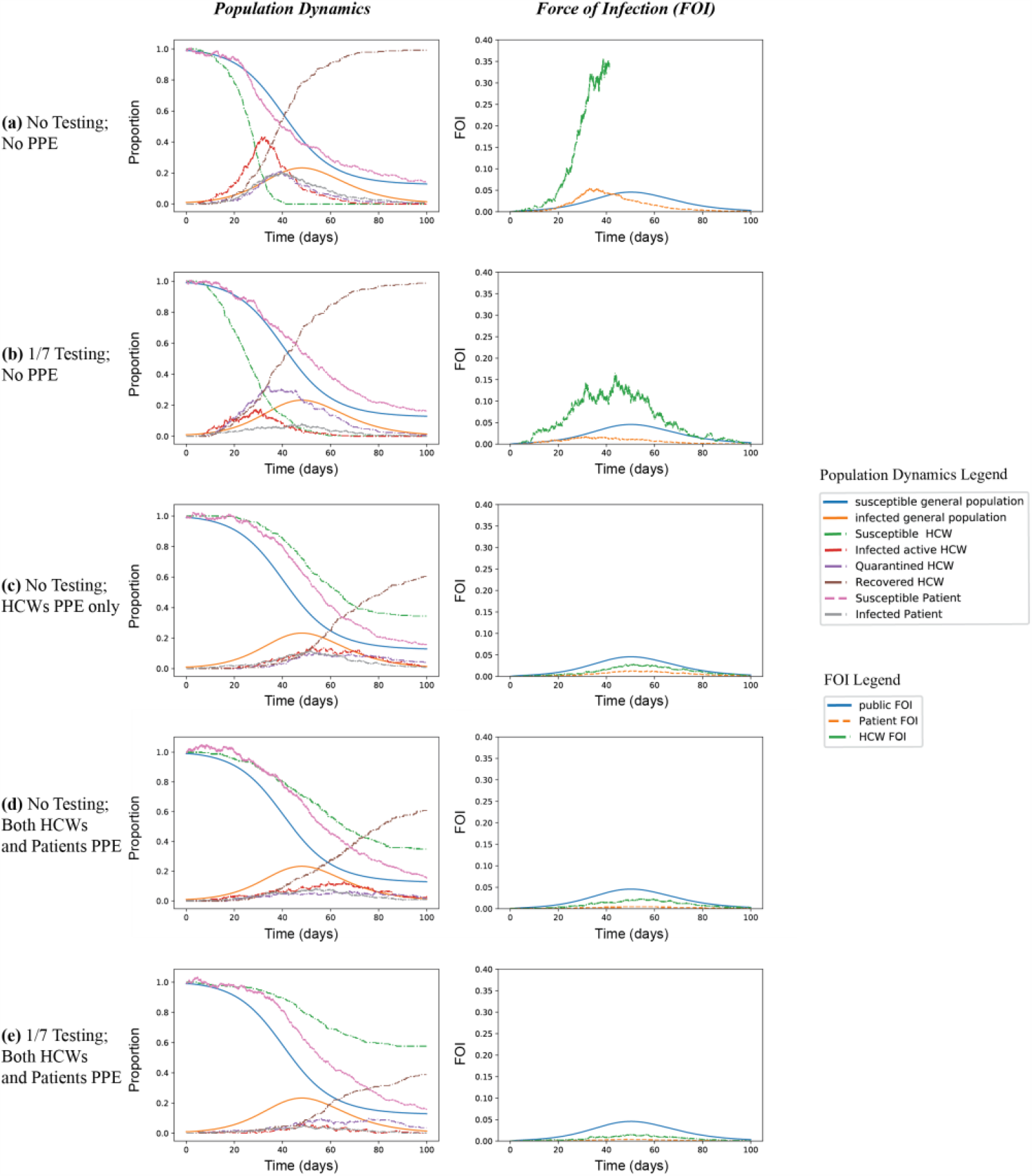
Comparing different scenarios of testing and PPE. Plots show the susceptible and infected portions of the cohort, and the force of infection. When there is no testing and no PPE (a), all HCWs are rapidly infected. The calculation of FOI on HCWs terminates once all have been infected. At peak about 40% of the HCWs are infected (not in quarantine), and shortly thereafter about 20% of the HCWs are under quarantine. The plots show that both testing (b) and PPE (c-d) can reduce the force of infection (FOI). But PPE has more substantial impacts on delaying and reducing the HCWs infection peak and the FOI. Noticeably, even only HCWs use PPE in (c), the infection peak and FOI in both HCWs and patients are reduced. (e) shows the impact of simultaneous use of testing and effective PPE.

### 2. Impacts of regular testing

Accurate virological testing is important to implement containment measures where a case is identified. HCWs have been recognized as an important group to receive testing both because of the exposure risks inherent in their profession and the potential consequences of their infection for others.^4^ Testing, especially while it has been scarce, has been understandably directed at those with symptoms. However, COVID-19 has a range of presentations, and infectious individuals may be currently asymptomatic.^17^ In the absence of testing, the asymptomatic patients and HCWs cannot be removed from the population and pose an infection risk to the rest of the cohort. We model this as a random testing rate of apparently uninfected patients and HCWs on a weekly basis *ω* = 1*/*7, we see a significantly lower force of infection (FOI) on both HCWs and patients (Figure 1(b)). It takes longer for the HCWs to all become infected, and the peak level of HCWs quarantine is higher as a result of more cases being identified. These impacts are expected to be larger for higher testing rates.

### 3. Impacts of PPE

PPE substantially delays the peak of infection and reduces FOI, even when only used by HCWs (Figure 1(c), (d), (e)). In many locations, PPE supply has been limited, leading to re-use of normally disposable items or in some improvised equipment. Our model also is concerned with PPE use throughout the non-COVID cohort, which implies use by staff who are not familiar with best practice for PPE use prior to the pandemic. So we investigated the impact of less effective PPE (whether due to improper use or lower quality equipment). we define perfect PPE as reducing the scaling factor of transmission to 1/8 of default values; Imperfect PPE is defined by the reduced effectiveness of PPE in preventing transmission, and can be considered to represent situations in which PPE shortages lead to diversion of supply to the COVID-19 cohort. For example, 50% effective PPE means that the use of PPE reduces the transmission rate by half. Based on the simulations (Figure 2), we find that even half effective PPE (Figure 2(b)) can bring down the FOI of HCWs near to that in the general population.

**Figure 2.**
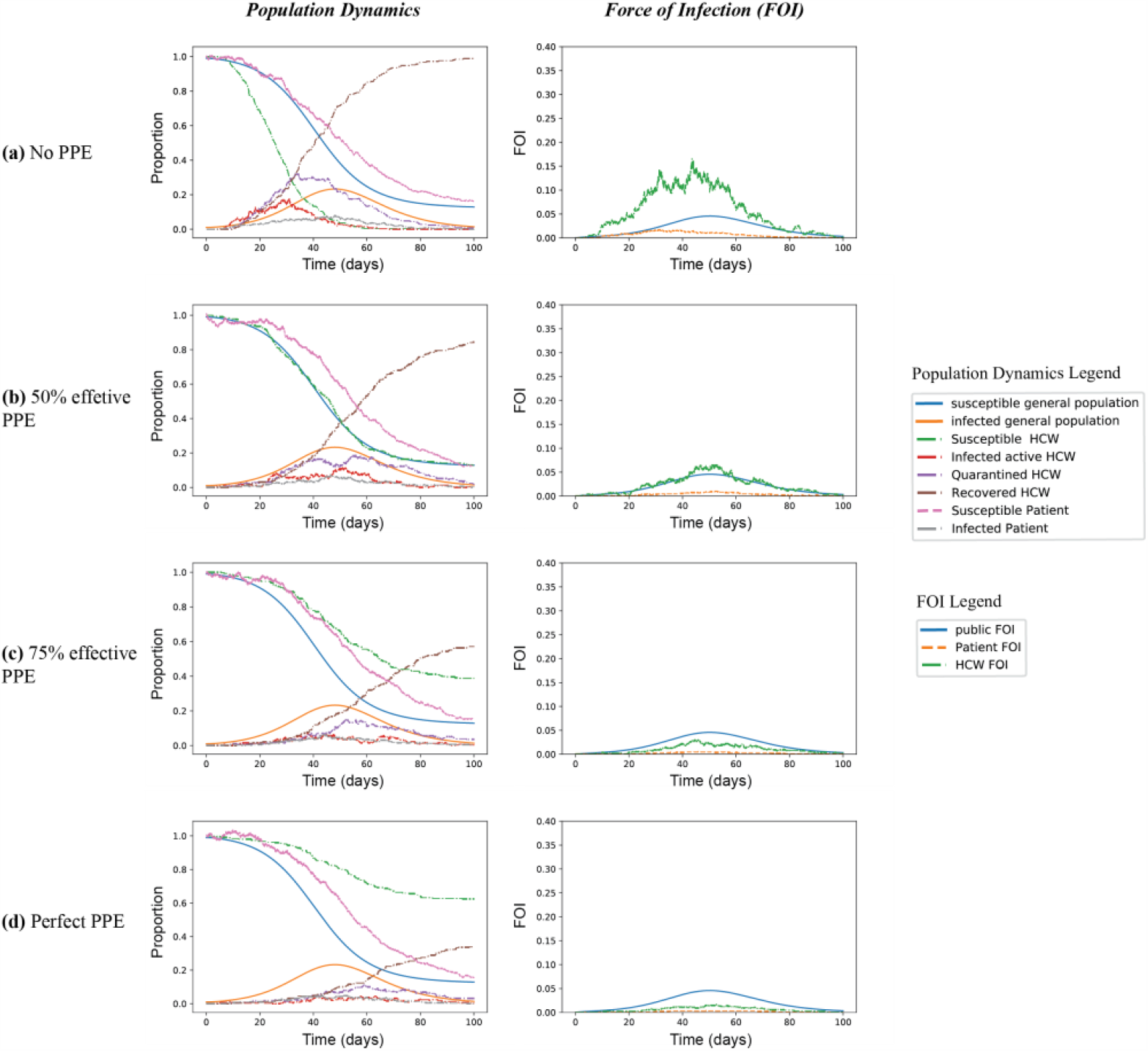
The relative protection of PPE in non-COVID-19 cohorts. In all scenarios testing rate *ω*=1/7 but with PPE of varying efficacy. At best we assume PPE reduces the scaling factors of nosocomial transmission to 1/8 of the default values (d); Plot (a) represents no PPE use; (b) represents prevention of 50% of transmission events; (c) represents prevention of 75% of transmission events. Results indicate that even modest reductions in transmission can reduce the force of infection inside healthcare setting.

### 4. Impacts of asymptomatic infection

The proportion of asymptomatic infections (18%–75%) varies among currently available epidemiological studies,^6,18–20^ we therefore examined scenarios in which varying proportions of infections were asymptomatic (q), in the presence of testing to detect them at rate *ω*= 1/7 (Figure 3). As shown, as the proportion of asymptomatic infections increases the FOI among HCWs increases with it, leading it to peak earlier with concomitant effects on patients (Figure 3(a-c)).

**Figure 3.**
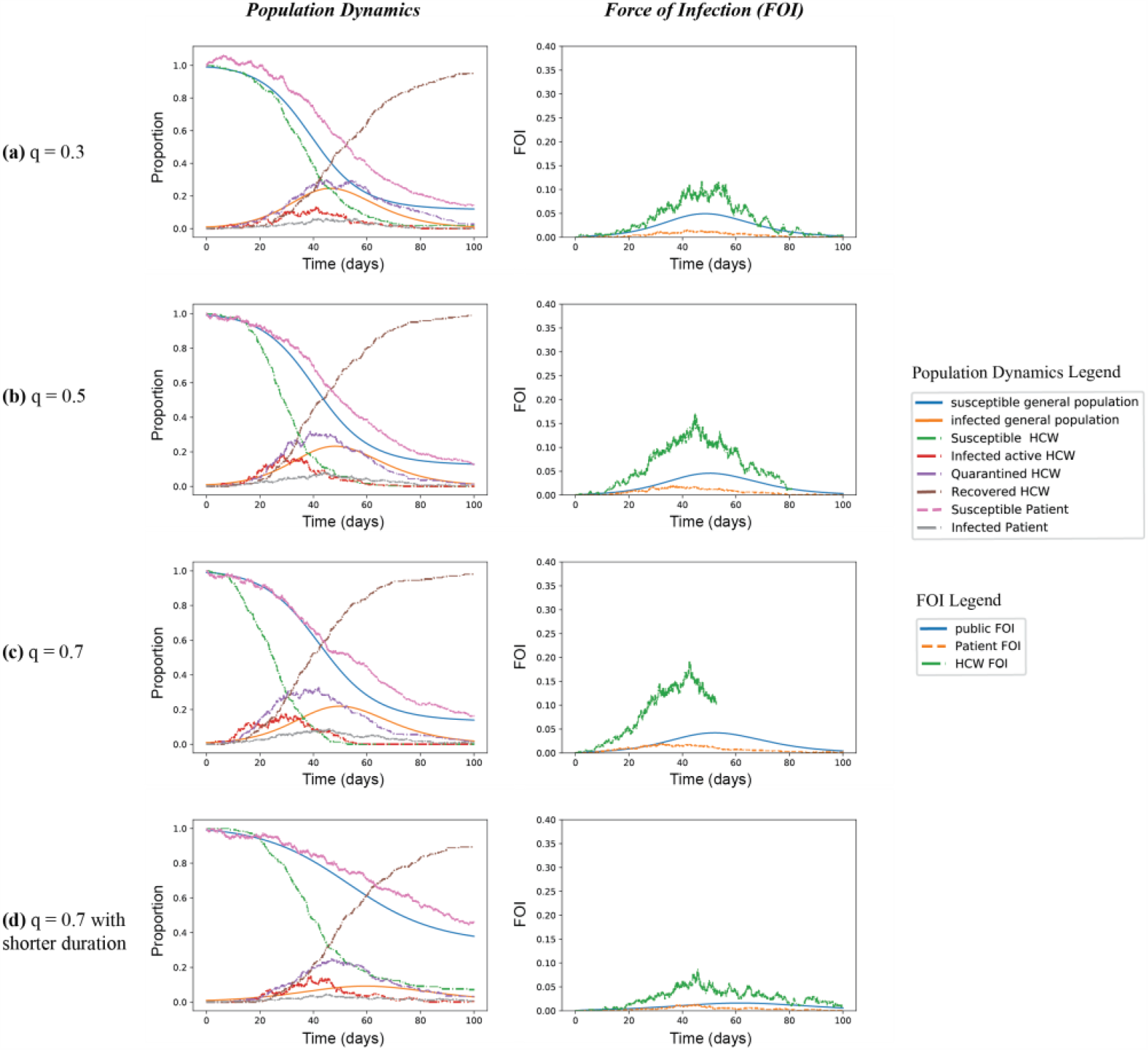
Comparison of scenarios with varying proportions of asymptomatic infections (q). Here all scenarios have a testing rate *ω*=1/7. The asymptomatic proportion was changed from the default value of q=0.5 in (b) to a lower value of q=0.3 in (a) and then to a higher value of q=0.7 in (c). To explore the impact of potential shorter duration of infectiousness of asymptomatic infections, the parameter of *γ*_*A*_ was changed from the default 1/9 to 1/5 with q=0.7 in (d). We find that the increasing proportions of asymptomatic infections can increase the peak of infected HCWs and patients, increase, the FOI, and reduce the peak of quanrantined HCWs. However, the duration of infectiousness of the asymptomatic has larger impacts, where under the higher proportion q=0.7, if the duration of infectiousness is shorter, the peak of infections and FOI can substantially reduce.

The effect of this is minor in comparison with the consequences of reducing the duration of the asymptomatic period (Figure 3(d)), which intuitively reduces the opportunity for exposure and transmission. This suggests the importance of testing for detecting asymptomatic or presymptomatic infected individuals among both HCWs and patients promptly. It also indicates the importance of PPE use among as many individuals as possible, in order to limit unwitting transmission from individuals not yet tested.

### 5. Impacts of cohort size

The probability that a given introduction establishes in a cohort is independent of *L* once *L* is reasonably large. However, the expected number of introductions is proportional to *L*. For this reason, the probability a cohort does not have a successful introduction increases as *L* decreases.

Assuming the introduction rate is proportional to the cohort size, the probability infection establishes itself into a cohort of size *L* is *e*_−*kL*_ for some *k* > 0. The value of *k* increases with the rate at which non-symptomatic infected individuals are admitted, the rate at which the general public transmits to patients or HCWs, and the transmission rate between individuals in the health-care system. The value of *k* decreases as the recovery rates and testing rates increase. The probability of at least one successful introduction into a cohort is thus 1−*e*_−*kL*_.

If infection is established within a cohort, it will typically infect some fraction of the total population. Like typical epidemics, this fraction is independent of the population size. So for larger populations, the number of infections increases.

This motivates the following observation: given a collection of cohorts that are small enough to each have a non-negligible chance of escaping infection, then joining them together increases the risk to all members of the cohorts. The cumulative distribution function of outbreak sizes for cohorts of different size is shown in Figure 4. This suggests that dividing into smaller cohorts and making efforts to minimize the risk of successful introduction can be an effective way to reduce risk of infection within the cohorts. Smaller cohorts also reduce the amount of additional testing required to identify secondary transmission among contacts once one case is identified.

**Figure 4.**
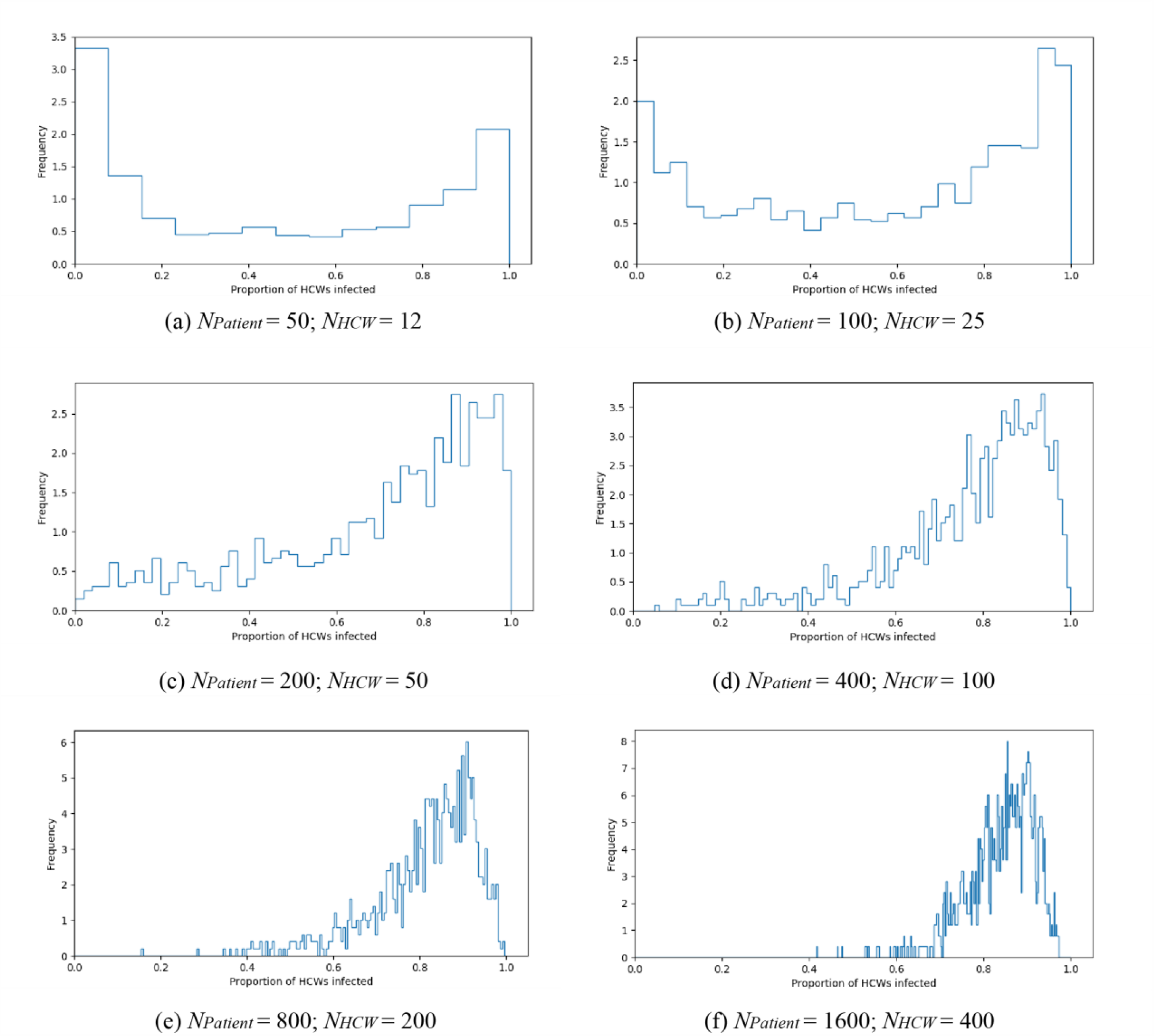
The impact of cohort size, based on 1000 simulations per cohort size. Probability density of outbreak sizes for different cohort sizes. As the cohort size increases the frequency of small outbreaks goes down. With large cohort sizes, all cohorts have outbreaks that infect a large fraction of people. With small cohort sizes, many cohorts have no outbreaks, or outbreaks that only infect a few.

Whether infection comes in through an externally infected HCW, a visitor, or an asymptomatic new infection does not significantly affect the outcomes. As long as the within cohort reproduction number (Appendix B) is greater than 1, once the infection is established in the cohort the dynamics will be dominated by the internal infection process.

## Discussion

COVID-19 presents an unprecedented challenge throughout all healthcare systems. The pronounced increases in the risk of severe disease or death that are found in older age groups, as well as patients suffering co-morbidities, demands that these at-risk groups be protected. And yet they are also disproportionately likely to require healthcare for conditions other than COVID-19. Contact and risk to the vulnerable can be reduced by innovations such as telemedicine consultations for chronic conditions, but urgent care will remain to be needed in acute cases. This work has been an attempt to evaluate the roles of cohorting, prompt and accurate diagnostic testing, and PPE in protecting patients and healthcare workers.

Our primary finding is that the relative impacts of interventions depend on the underlying properties of the disease and in particular infection from currently asymptomatic individuals. The possibility of this has been apparent for some time and it has recently been confirmed to be responsible for a large fraction of transmission events.^17,21^ We find that this makes little impact on the force of infection (with the caveat that it depends on the duration of the asymptomatic period), but it magnifies the impact of effective PPE. The potential for transmission from pre-symptomatic individuals has long been known to be a crucial component in how hard we expect it to be to control an infectious disease.^22^ This model confirms that if we wish to prevent SARS-CoV-2 transmission among the vulnerable noncovid cohort all individuals should be assumed to be infectious, both staff and patients. Where appropriate PPE is available it is being widely used throughout healthcare, and indeed use of cloth masks is now recommended for the general public by the Centers for Disease Control. However ample PPE may not continue to be available in all settings, and PPE for the non-COVID-19 cohort is an important element of planning. Notably, the impact of weekly random testing of staff and patients in the non-COVID cohort is unable to prevent infection becoming established in the absence of other interventions. These findings are also relevant to developing countries where testing may not be widely available, or anywhere a tradeoff exists between testing and PPE.

Our findings regarding the size of sub cohorts are somewhat nuanced, but identifying some general trends is straightforward; reducing any of the transmission rates is unsurprisingly important in reducing transmission. However, by keeping subcohorts smaller, we reduce the probability that infection establishes. Further, by reducing interaction amongst HCWs, we can reduce transmission risks. If infection reaches a cohort, the introduction may fail to establish itself. However, modeling shows that when an infection does establish it tends to have an increased early growth rate.^23^ Mathematically this can be interpreted as a consequence of the fact that if on average a small outbreak would grow by a factor of R_0_ at each generation, but some go extinct, then those that do not go extinct must have increased transmissibility in order to achieve the observed dynamics.^24^ This means that interventions that increase the probability of causing 0 transmissions from an introduction are of particular importance. In the presence of a very high force of infection from the community at large they are of limited value. However, in combination with a sustained effort to prevent the introduction of infections (and at the initial stage of the pandemic) smaller cohorts in which HCW are kept separate may have value in preventing establishment of the infection in the healthcare setting. Moreover, this work does not model any attempted mitigation strategies in the community at large to reduce the force of infection. In the presence of community mitigation strategies, the value of subcohorting is expected to be enhanced.

While cohorting is understood to be important,^25^ subcohorting has so far received less attention. Our findings suggest this could be an important strategy, especially in combination with community mitigation strategies and in settings where PPE and testing may be in short supply.

There are several important elements of the COVID-19 pandemic and SARS-CoV-2 biology that are not captured by our model. We have assumed an unmitigated outbreak outside the non-COVID-19 cohort, which is not the case in most locations. However, much of the most important dynamics we observe happen early on, and so our findings will be relevant independent of the details of the pandemic outside. We also do not directly model the consequences of transmission in the health care setting; obviously transmission to elderly patients or otherwise vulnerable individuals is expected to have an outsize impact on overall mortality and the strain on healthcare in general. We have also not considered the consequences of an overdistributed *R*0. The SARS-CoV-1 outbreak, as well as MERS outbreaks have both been characterised by superspreading events in healthcare settings,^26,27^ and future work should explicitly consider the impact of these. This is especially relevant to our findings on the size of individual subcohorts, because it is known that an overdispersed *R*0 can lead to situations in which most disease introductions go extinct.^24^ Finally we have assumed that testing is accurate, while in reality sensitivity likely depends on the stage of infection, being highest early on and declining.

In reality we might wish to distinguish between truly asymptomatic and presymptomatic cases and those in which the presentation of disease is unusual and so the infection is not suspected. The overall impact however is similar for our model. We have not modeled the potential for transmission to be further curtailed by aggressive isolation of contacts of known cases, either among patients or HCWs. We have also not modeled the impact of strategies in the community to limit transmission and infection, such as physical distancing or salutary sheltering, instead assuming a simple and symmetrical force of infection. These are not likely to alter our major conclusions. Indeed as we note, mitigation to decrease the FOI in the community would likely increase the beneficial effects of subcohorting strategies.

As communities around the globe confront the pandemic, the most important way to reduce transmission in healthcare settings is to ensure an adequate supply of PPE to reduce transmission. Testing should also be made available both to identify those who are infected and those who have been infected, and innovative approaches will need to be taken to minimize the pandemic threat. Subcohorting within institutions is a simple and potentially underutilized approach that could also help reduce healthcare transmission, especially in lower incidence settings and in combination with strategies to mitigate the pandemic in the community at large. We hope that our analysis will motivate future action to preserve lives.

## Data Availability

The analysis is based on model construction and simulations. The specific details of our simulations are described in the Appendix. The simulations are scripted in Python  and available to the open public.

https://github.com/joelmiller/HospitalCOVID19

## Acknowledgment

XQ and WPH were supported by the National Institute of General Medical Sciences of the National Institutes of Health under Award Number U54GM088558. JCM was supported by startup funding from La Trobe University. The content is solely the responsibility of the authors and does not necessarily represent the official views of the National Institutes of Health.

## Author contributions

JCM conceived of the study, conducted the analysis, interpreted results, wrote and edited the manuscript. XQ searched literature, conducted the analysis, interpreted results, wrote and edited the manuscript. DM searched literature, wrote and edited the manuscript. WPH conceived of the study, interpreted results, wrote and edited the manuscript.

## Appendix

### A Model description

#### A.1 Dynamics in general public

We assume that the infection status of individuals in the general public can be separated into 6 classes: Individuals begin Susceptible (*S*). After receiving a transmission, they become Exposed (*E*) but not infectious. They then become Infectious but not symptomatic (presymptomatic infectious) *I*_1_, Infectious and symptomatic *I*_2_, and recovered *R* (Figure A.1). There is considerable dispute about the distinction between truly asymptomatic infection, presymtomatic or subclinical and so on. Here we use this term to capture those who could not be reasonably expected to alter their behavior to avoid infecting others.

**Figure A1:**
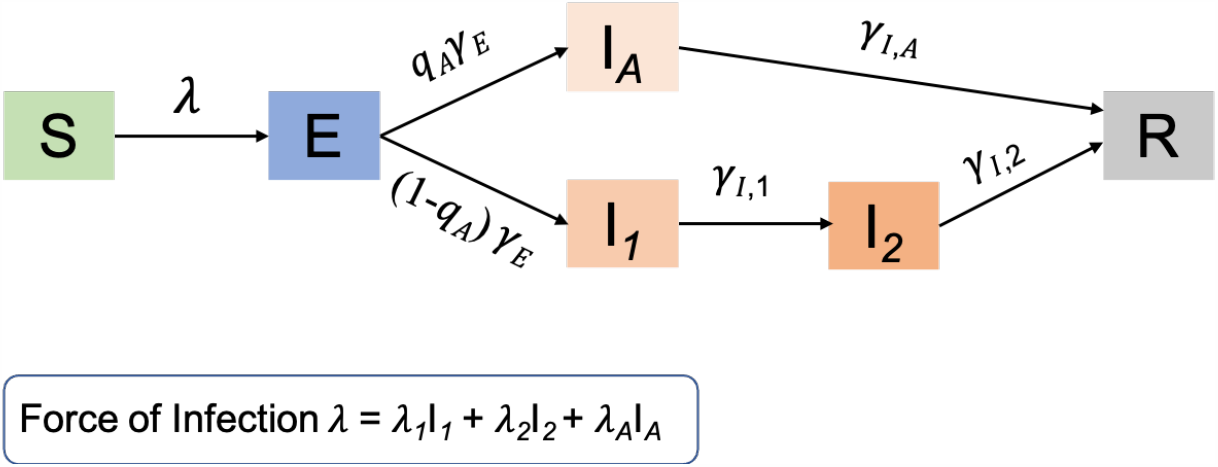
Compartmental model of the dynamics in general public

We assume deterministic dynamics in the general population as, at least initially, we expect the epidemic to be more advanced and the incidence to be higher outside the healthcare setting. We have

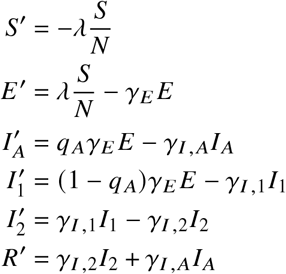

where *λ* = *λ*_1_*I*_1_ + *λ*_2_*I*_2_ + *λ*_*A*_*I*_*A*_.

#### A.2 Dynamics in cohort of (presumed) uninfected patients

We now derive the equations which will be used in patients. These will calculate the rates at which transitions happen.

We assume if a patient becomes symptomatic or is identified as infected through testing, the patient is immediately removed from the cohort, so we track *S*_*P*_, *E*_*P*_, *I*_*A*_, and *I*_1,*P*_ (Figure A.2).

New patients arrive at rate *b*. We assume that the arriving patients are chosen randomly from the non-symptomatic population. The probability that an non-symptomatic, but infected individual enters the population is reduced by a factor *ρ*. This means

**Figure A2:**
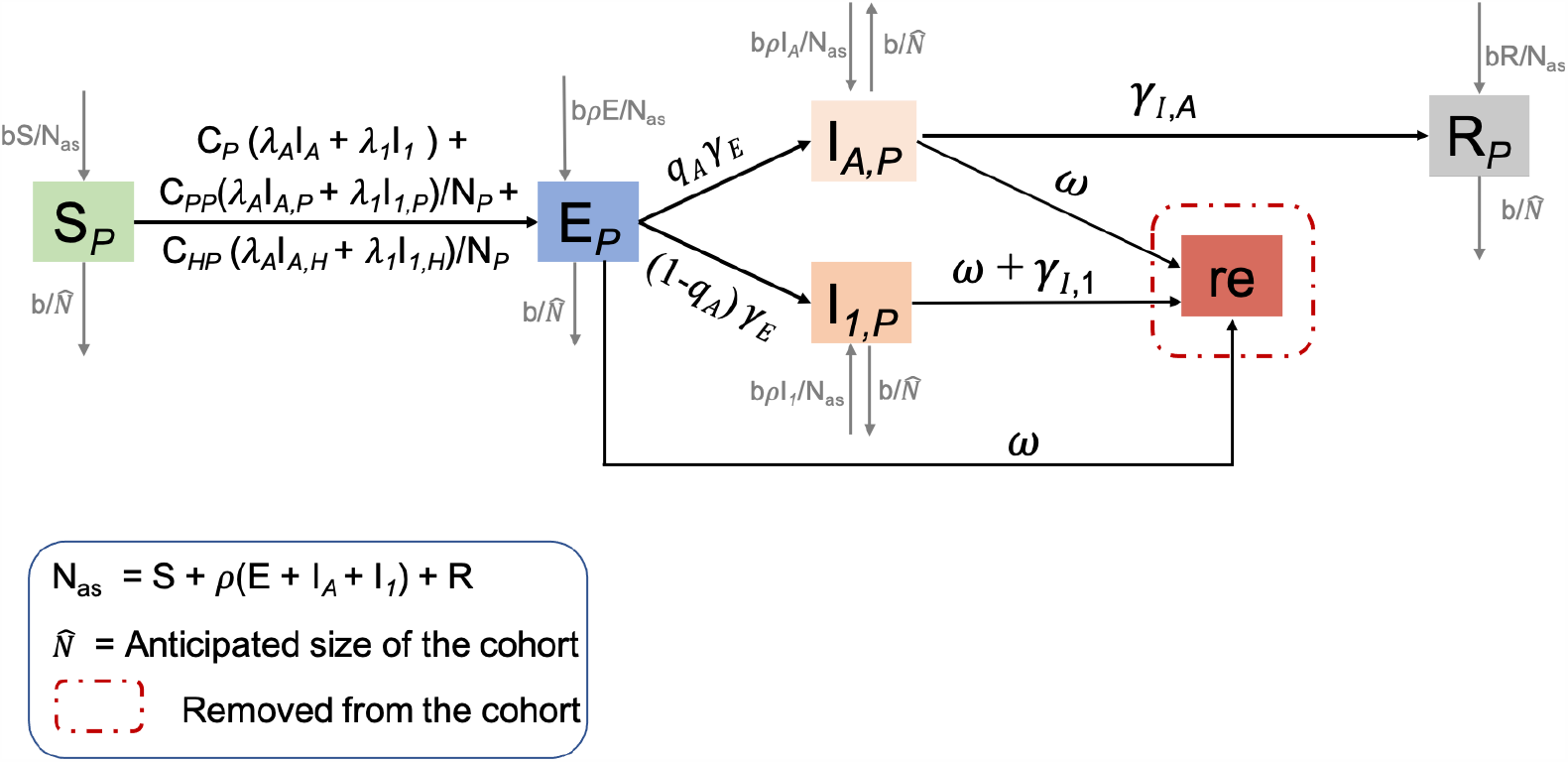
Compartmental model of the dynamics in non-COVID-19 patients factor *ρ*. This means

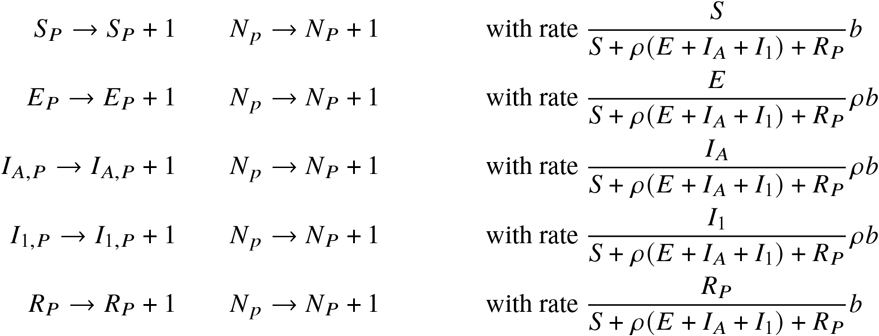

We also have departures from the population (deaths or discharge) at combined rate 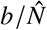 where 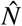 is the anticipated size of the cohort. So all of the variables for the patients (*S*_*P*_, *E*_*P*_, etc.) all reduce by 1 independently as Poisson processes with rate 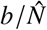 times their current value (so for example, *S*_*P*_→ *S*_*P*_− 1 and *N*_*P*_ →*N*_*P*_ −1 at rate 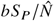).

We assume there are several potential sources of infection for a susceptible patient:

- asymptomatic/presymptomatic infections in the general population, with rate *C*_*P*_ (,*λ*_*A*_*I*_*A*_ +, *λ*_1_*I*_1_).
- Infections from other asymptomatic or presymptomatic patients with rate

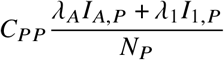

where *N*_*P*_ = *S*_*P*_ + *E*_*P*_ + *I*_1,*P*_ + *I*_*A,P*_ + *R*_*A,P*_ is the number of patients in the cohort.
- Infections from asymptomatic or presymptomatic HCWs, with rate

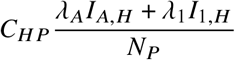

So we find

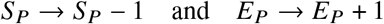

at rate

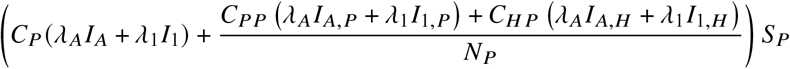

Exposed individuals transition to infectious at rate *γ*_*E*_, so

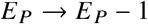

at rate *γ*_*E*_ *E*_*P*_. Of those that transition, a fraction *q*_*A*_ become asymptomatic and (1 − *q*_*A*_) become presymptomatic. So when this transition happens, with probability *q*_*A*_ we have *I*_*A,P*_ → *I*_*A,P*_ + 1 and with probability (1 −*q*_*A*_) we have *I*_1,*P*_ → *I*_1,*P*_ + 1

Due to testing, infected patients can be removed from the cohort with rate *ω*. This gives:

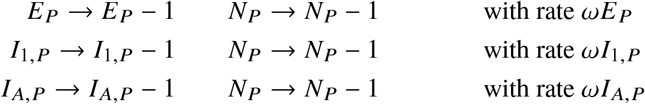

Finally infectious presymptomatic individuals transition to symptomatic and are immediately removed at rate *γ*_*l*, 1_, so

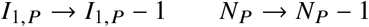

at rate *γ*_*l*, 1_*I*_1,*P*_.

#### A.3 Dynamics in HCWs

Note *N*_*H*_ = *S*_*H*_ + *E*_*H*_ + *I*_1,*H*_ + *I*_*A,H*_ + *R*_*H*_, so it does not include quarantined HCWs (Figure A.3).

**Figure A3:**
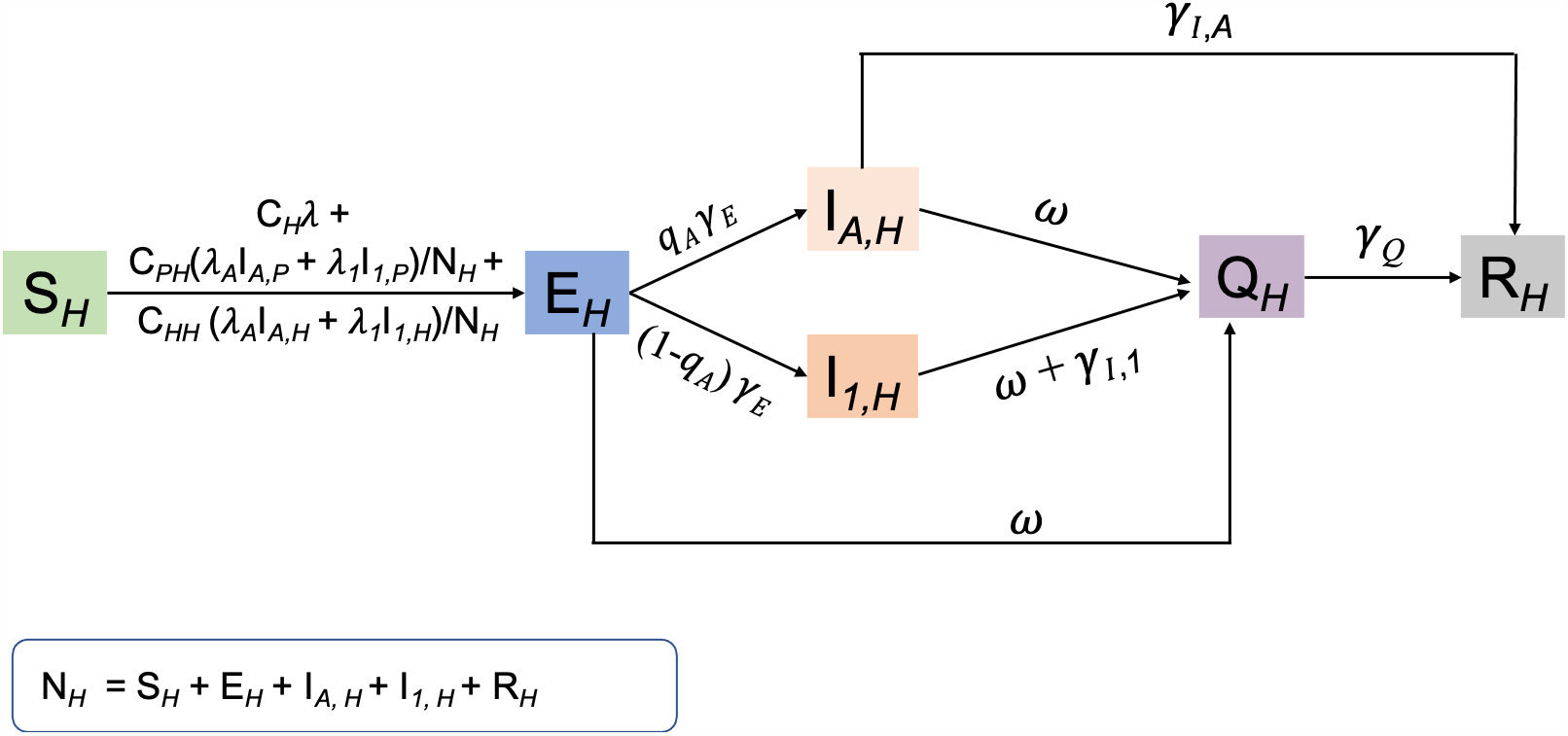
Compartmental model of the dynamics in healthcare workers

We assume that the HCWs in the cohort suffer a force of infection

- from the general public, with rate *C*_*H*_ (*λ*_1_*I*_1_ + *λ*_2_*I*_2_ + *λ*_*A*_*I*_*A*_) = *C*_*H*_, *λ* where *C*_*H*_ < 1.
- a force of infection from their patients.

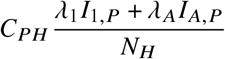

where *N*_*H*_ is the number of active HCWs and *C*_*PH*_ is a factor scaling the patient to HCW transmission rate relative to the patient to patient transmission rate.
- They also experience a force of infection from fellow HCWs

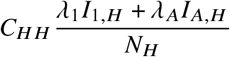

where *C*_*H H*_ scales the HCW-HCW transmission rate.

So we find

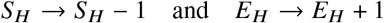

with rate

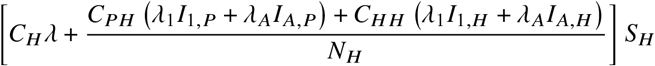

Exposed individuals transition to infectious at rate *γ*_*E*_. With probability_*A*_ that transition is to an asymptomatic state, so

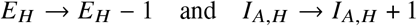

at rate *q γ*_*E*_ *E*_*H*_. With probability 1 − *q*_*A*_ the transition is to the presymptomatic state, so

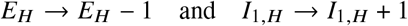

with rate (1−*q*) *γ*_*E*_ *E*_*H*_.

Testing occurs with rate *ω*. Identified individuals are quarantined, temporarily removing them from the active population

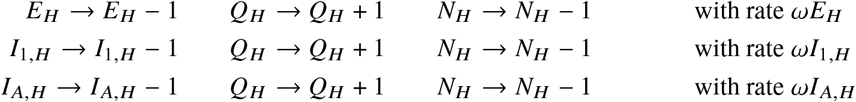

Infectious presymptomatic HCWs transition to symptomatic *γ*_*l*,1_. These individuals are quarantined and do not interact with others. So this transition reduces *N*_*H*_ by 1. So

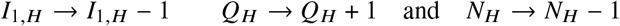

at rate *γ*_*l*, 1_*I*_1,*H*_.

However, they recover and return to the population at rate *γ*_*Q*_. This transition increases *N*_*H*_ by 1:

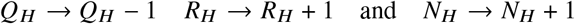

The asymptomatic HCWs recover with rate *γ*_*l, A*_, so

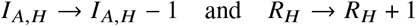

at rate *γ*_*l, A*_*I*_*A,H*_

### B Reproduction number within cohort

To calculate the reproduction number within the cohort, we need to calculate the probability that an infected patient or HCW enters each infected state and the expected number of infections within the cohort that result. We start by looking at a newly infected patient. Note that the probability an infected individual becomes asymptomatically infected before being tested or discharged is 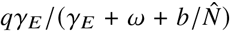 and the probability of becoming presymptomatic is 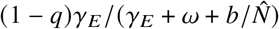.

If the patient develops asymptomatic infection, the expected number of new patients infected will be *C*_*PP*_, *λ*_*A*_/(*γ*_*A*_+ *ω*). If the patient enters the presymptomatic state, the expected number of new patients infected before being tested or discharged will be 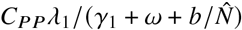. The expected number of transmissions to other patients is thus

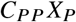

Where

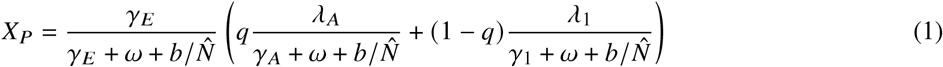

The expected number of transmissions to HCWs from an infected patient is *C*_*PH*_ *X*. Similarly the expected number of transmissions from an infected HCW is *C*_*H P*_ *X*_*H*_ to a patient and *C*_*H H*_ *X*_*H*_ to an HCW where *X*_*H*_ is the same as *X*_*P*_, but with 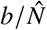 replaced by 0.

So we can capture the number of infections in the “next generation” by the matrix

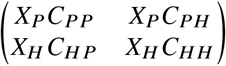

The dominant eigenvalue of this matrix is the reproduction number

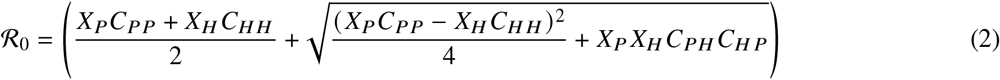

The expression for *X*_*H*_ and *X*_*P*_ suggest increasing the testing rate *ω* would be a particularly important step for reducing *R*_0_ because it appears in the denominator of the first term and in the denominators of both terms inside the parentheses in equation (1). Similarly, any interventions that simultaneously impact both infectiousness of infected individuals and susceptibility of susceptible individuals will also have significant impact.

## C Limitations and additional comments

There are some particularly important effects which we have not included in the model:

- We assume that testing continues at a constant rate in the cohort. This neglects an enhanced level of testing that might be expected if an infection is detected.
- We ignore heterogeneity in infectiousness. In the general population, this is not significant. However, as seen in [1, 2], this can have an important impact on the establishment of a disease following introduction. Higher heterogeneity makes diseases less likely to establish, but those that successfuly establish have higher average initial growth than predicted from *R*_0_.
- We have neglected HCWs transmission from interactions with those involved in the COVID-19 cohort or other non-COVID-19 cohorts.
- We assume that all patients and HCWs are monitored closely enough that individuals are immediately identified once symptomatic.

